# NutrIA: Development and Internal Validation of a Hybrid Clinical Decision Support System for Personalized Preventive Nutrition

**DOI:** 10.64898/2026.08.13.26359723

**Authors:** Bohdan Isaiev, Iryna Stukalova

## Abstract

**Background:** The growing burden of lifestyle-related chronic diseases has increased the need for clinically interpretable decision-support tools capable of integrating artificial intelligence with evidence-based preventive nutrition. Although machine learning has shown considerable potential for health risk prediction, most existing approaches remain limited to isolated predictive models or conventional nutritional software, with little integration of multidimensional clinical assessment and personalized recommendations.

**Objective:** To develop and internally validate NutrIA, a hybrid web-based Clinical Decision Support System (CDSS) that combines machine learning, validated clinical assessment, structured clinical reasoning and personalized nutritional recommendations for preventive medicine.

**Methods:** NutrIA was developed using harmonized data from the National Health and Nutrition Examination Survey (NHANES, 1988–2018). A supervised machine learning model was trained to estimate 5-, 10- and 20-year all-cause mortality risk and subsequently integrated with an adaptive clinical questionnaire, validated screening instruments, nutritional indicators, dietary clustering, clinical phenotyping and a transparent rule-based recommendation engine within a unified web-based platform.

**Results:** The predictive model achieved ROC-AUC values of 0.894, 0.914 and 0.923 for 5-, 10- and 20-year mortality prediction, respectively. The implemented CDSS incorporates an adaptive questionnaire (151 items), 39 validated clinical assessment instruments, 17 clinical phenotypes and 31 dietary clustering modules to generate individualized nutritional and lifestyle recommendations together with an automated clinical report. The integrated framework translates probabilistic risk estimates into clinically interpretable decision support for personalized preventive nutrition.

**Conclusions:** NutrIA demonstrates the technical feasibility of integrating machine learning with knowledge-based clinical reasoning within a single web-based CDSS for preventive nutrition. Although external validation and prospective clinical evaluation are required before routine implementation, the proposed architecture represents a promising step toward clinically interpretable artificial intelligence for personalized nutritional care.

## 1. Introduction

Chronic non-communicable diseases (NCDs), including obesity, type 2 diabetes mellitus, cardiovascular diseases and other cardiometabolic disorders, remain the leading cause of morbidity and mortality worldwide. Their increasing prevalence represents one of the greatest public health challenges of the twenty-first century and highlights the need for effective preventive strategies capable of identifying individuals at increased risk before clinical manifestations occur [1,2]. Since many of these conditions are strongly influenced by modifiable factors such as dietary habits, lifestyle, body composition and metabolic status, preventive nutrition has become a fundamental component of modern healthcare. In this context, personalized nutrition has emerged as a promising approach to improve disease prevention by tailoring dietary interventions to the characteristics and risk profile of each individual [3,4].

Recent advances in artificial intelligence (AI) and machine learning (ML) have accelerated the development of computational tools capable of analysing large and heterogeneous health datasets to identify complex relationships that are difficult to detect using conventional statistical methods [5–7]. Within nutrition and preventive medicine, these technologies have been applied to dietary assessment, prediction of chronic diseases, food recommendation systems and clinical risk estimation [8–12]. In parallel, Clinical Decision Support Systems (CDSSs) have become increasingly relevant by assisting healthcare professionals in integrating patient information with scientific evidence to support clinical decision-making [13,14].

Despite these advances, current digital solutions remain highly fragmented. Commercial nutrition platforms such as Nutrium, Dietbox and Healthie facilitate patient management, dietary assessment and nutritional follow-up but do not incorporate predictive models capable of estimating future health risks from multidimensional clinical information. Conversely, validated clinical calculators such as QRISK3 and FINDRISC provide estimates for specific diseases, including cardiovascular disease or type 2 diabetes, but are restricted to a single clinical outcome and do not integrate comprehensive nutritional assessment or personalized dietary recommendations [15,16]. Similarly, many AI-based predictive models reported in the literature primarily focus on maximizing predictive performance while offering limited clinical interpretability and lacking integration into complete decision support systems applicable in routine clinical practice [6,14,17–19].

These limitations are particularly relevant in preventive nutrition, where clinical decision-making requires the simultaneous interpretation of demographic, anthropometric, biochemical, dietary, behavioural and lifestyle information [4,7,8, 10]. Existing approaches rarely integrate these heterogeneous data sources with validated nutritional indices, evidence-based clinical reasoning and individualized dietary recommendations within a unified clinical decision support framework [10, 13,17]. Consequently, healthcare professionals frequently rely on multiple independent tools, reducing workflow efficiency and limiting the implementation of truly personalized preventive interventions [13,18].

To address these limitations, this study presents NutrIA, a hybrid web-based CDSS for personalized nutrition and preventive medicine. NutrIA integrates supervised machine learning models developed using data from the National Health and Nutrition Examination Survey (NHANES) [20], validated clinical assessment tools, an adaptive questionnaire, automated risk stratification and a rule-based recommendation engine to generate individualized dietary recommendations and clinical reports. By combining predictive analytics with structured clinical reasoning, NutrIA provides an interoperable framework for evidence-based nutritional decision support.

The aim of this study was to develop and internally validate a hybrid web-based CDSS integrating machine learning and knowledge-based reasoning to support personalized nutrition and preventive medicine using NHANES data [20].

## 2. Materials and Methods

### 2.1. Study design

This study describes the development and internal validation of NutrIA, a hybrid web-based Clinical Decision Support System (CDSS) for personalized nutritional assessment and preventive medicine. The development process comprised two complementary components. First, supervised machine learning models were developed using data from the National Health and Nutrition Examination Survey (NHANES) to estimate long-term mortality risk. Second, these models were integrated into a knowledge-based clinical decision support framework incorporating validated assessment tools, an adaptive questionnaire, automated risk stratification and personalized dietary recommendations. The resulting system was implemented as a web-based platform to support evidence-based nutritional decision-making.

The study followed a structured workflow including data acquisition and preprocessing, predictive model development and internal validation, integration of clinical knowledge and implementation of the web-based platform. The overall methodology is summarized in **Figure 1**.

**Figure 1.**
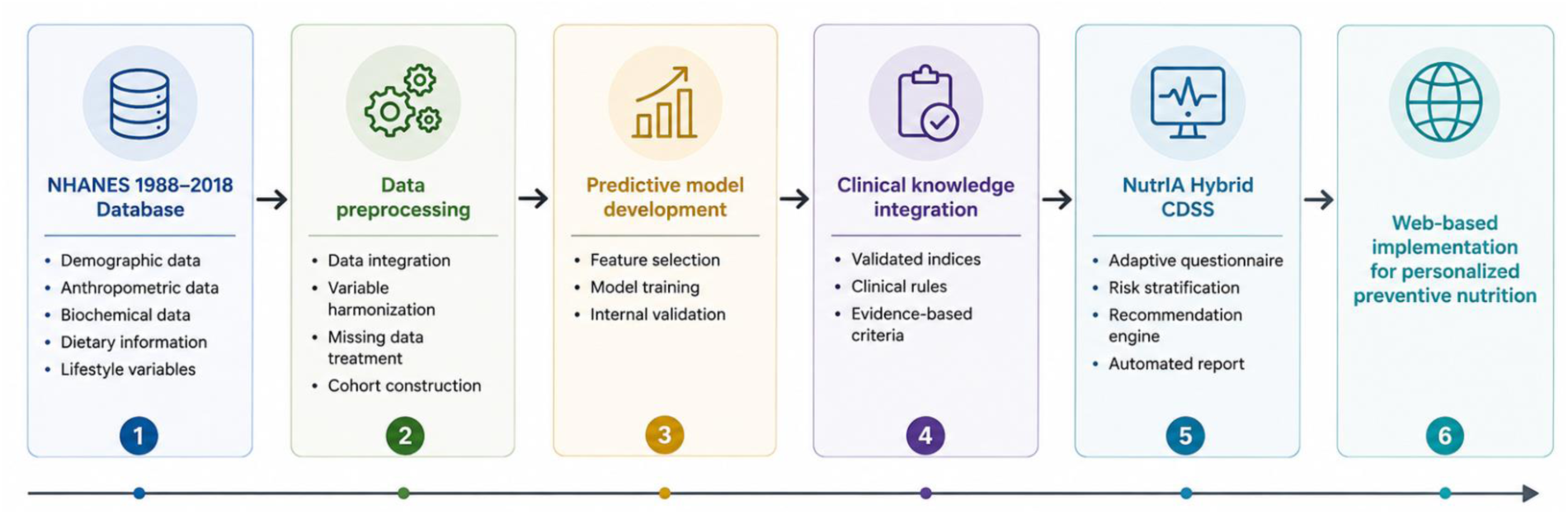
Overall methodological workflow of the NutrIA development process. The workflow comprises data acquisition and preprocessing, predictive model development, knowledge-based system integration and implementation of the web-based Clinical Decision Support System.

### 2.2. Data source

The predictive model underlying NutrIA was developed using data from the National Health and Nutrition Examination Survey (NHANES), a nationally representative health survey conducted by the National Center for Health Statistics (NCHS) of the Centers for Disease Control and Prevention (CDC) [20]. Publicly available data from NHANES III (1988–1994) together with the continuous NHANES cycles conducted between 1999 and 2018 were used in this study. To facilitate longitudinal analyses across multiple survey cycles, a harmonized research database was employed [21].

NHANES combines household interviews, standardized physical examinations, laboratory analyses, dietary assessments and health questionnaires to provide a multidimensional characterization of the health status of the non-institutionalized civilian population of the United States. Across the study period, the integrated database contained information from more than 130,000 participants, representing one of the most comprehensive publicly available epidemiological resources for population health research. The survey adopts a complex multistage probability sampling design and provides sampling weights that enable nationally representative estimates and support the development of robust predictive models.

The selection of NHANES over other publicly available epidemiological datasets was based on three main considerations. First, its large sample size and nationally representative design provide substantial statistical power and improve the generalizability of predictive models. Second, the availability of linked mortality data through the National Death Index (NDI) Linked Mortality Files [22] enables long-term follow-up and the development of mortality prediction models at multiple time horizons. Third, NHANES integrates demographic, anthropometric, biochemical, dietary, behavioral and clinical information within a single standardized framework, allowing the simultaneous evaluation of multiple determinants of health and nutrition.

The analytical database included demographic, anthropometric, laboratory, dietary, health questionnaire, medication and occupational datasets. Together, these modules provided multidimensional information on socioeconomic status, nutritional intake, lifestyle behaviours, chronic diseases and biochemical markers required for predictive model development. The principal data modules incorporated into NutrIA are summarized in **Table 1**.

**Table 1.**
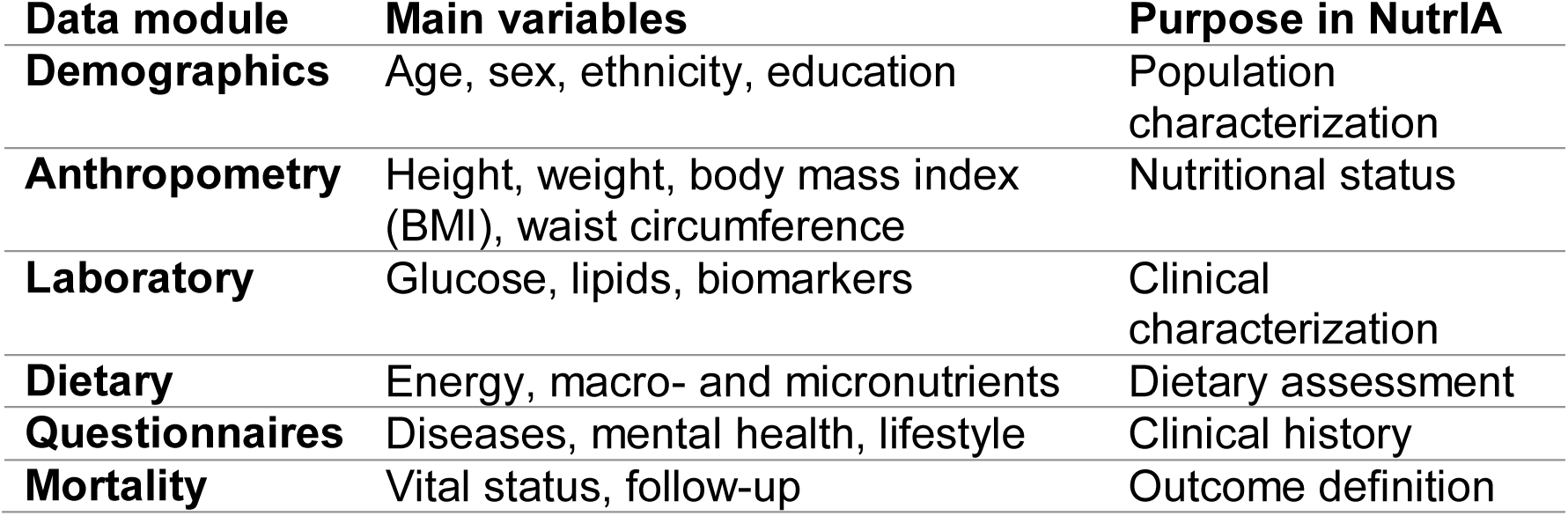
NHANES data modules included in the development of NutrIA.

Mortality outcomes were obtained from the NHANES Linked Mortality Files through linkage with the National Death Index (NDI) [22]. These files include information on vital status and follow-up time, allowing the association of baseline clinical and nutritional characteristics with long-term mortality outcomes. Although the complete NHANES database contains information on more than 130,000 participants, mortality information is available only for individuals successfully linked to the National Death Index, resulting in a smaller subset eligible for predictive modelling.

### 2.3. Data preprocessing

Prior to predictive modelling, the integrated NHANES dataset underwent a comprehensive preprocessing pipeline to ensure data consistency, quality and suitability for machine learning analyses. Given that NHANES data were collected over multiple survey cycles spanning more than three decades, substantial harmonization procedures were required before constructing the final analytical dataset.

#### 2.3.1. Data integration and cleaning

NHANES data are distributed across multiple survey cycles and thematic modules containing demographic, anthropometric, dietary, laboratory and clinical information. Individual records were linked using the participant identifier (SEQN) and survey cycle identifier (SDDSRVYR), resulting in a unified analytical dataset.

Variables were harmonized across survey cycles by standardizing variable names, recoding categorical variables, converting measurement units where necessary and removing duplicated variables introduced by methodological changes between NHANES cycles.

A systematic data cleaning procedure was performed before model development. Non-informative variables, including empty columns, technical identifiers and administrative variables without clinical relevance, were removed. Clinically plausible extreme observations were retained to preserve population variability, whereas inconsistent or clearly erroneous values were excluded according to predefined quality control criteria.

Variables with excessive missingness were excluded. For the remaining variables, missing values were imputed using the median for continuous variables and the mode for categorical variables when required for model development.

#### 2.3.2. Construction of the analytical cohort

Following preprocessing, an analytical cohort was constructed by applying predefined eligibility criteria. Participants aged ≥18 years with complete demographic information and valid mortality follow-up were included in the analytical cohort.

After preprocessing and eligibility selection, the final analytical dataset comprised 59,064 participants and 759 variables. The evolution of the dataset during preprocessing is summarized in **Table 2**.

**Table 2.** Dataset evolution during preprocessing.

| Stage | Participants | Variables |
| --- | --- | --- |
| Initial integrated NHANES dataset | 136,753 | 4,362 |
| After data integration, harmonization and cleaning | 101,316 | 1,364 |
| Final analytical cohort | 59,064 | 759 |

### 2.4. Development of the predictive model

#### 2.4.1. Outcome definition

The primary objective of the predictive model was to estimate the probability of all-cause mortality at clinically relevant time horizons of 5, 10 and 20 years using demographic, anthropometric, socioeconomic, lifestyle and self-reported clinical variables derived from NHANES. Mortality was selected as the primary outcome because it represents a robust and objective endpoint that integrates the cumulative effects of multiple health determinants and is widely used in preventive medicine and risk prediction research.

Mortality outcomes were obtained from the NHANES Linked Mortality Files as binary event indicators with corresponding follow-up time from baseline examination until death or censoring.

To facilitate clinical interpretation and implementation within the NutrIA Clinical Decision Support System (CDSS), three independent binary classification tasks were defined corresponding to mortality within 5, 10 and 20 years after baseline assessment. For each prediction horizon, participants who died within the specified follow-up period were classified as positive cases, whereas individuals remaining alive beyond the corresponding time horizon were classified as non-events. Participants without sufficient follow-up to determine their status within a given prediction window were excluded from the corresponding analysis in order to avoid outcome misclassification.

Fixed prediction horizons of 5, 10 and 20 years were selected to facilitate clinical interpretation and implementation within the NutrIA CDSS.

#### 2.4.2. Predictor selection

Following data preprocessing, the analytical dataset comprised 759 candidate variables representing multiple domains of health, nutrition and lifestyle. Given the high dimensionality of the dataset, an additional predictor selection procedure was performed to identify variables with the greatest predictive relevance while maintaining model interpretability and facilitating future implementation within a clinical decision support system.

Predictor selection followed a hybrid strategy combining data-driven feature importance analysis with expert-guided clinical selection. Preliminary predictive models were developed using the complete set of available variables. The relative contribution of each predictor was then evaluated using permutation feature importance, which quantifies the decrease in model performance after randomly permuting individual variables [23]. Variables demonstrating stable contributions across the three prediction horizons and showing consistency with current epidemiological evidence were prioritized for further consideration.

Final predictor selection considered clinical relevance, epidemiological evidence, feasibility for self-reported assessment, interpretability and applicability within the NutrIA CDSS. Variables showing excessive collinearity, limited clinical utility or substantial missingness were excluded. The final predictor set included variables from five principal domains, as summarized in **Table 3**.

**Table 3:** Predictor domains included in the final operational model.

| Domain | Variables included | Rationale for inclusion |
| --- | --- | --- |
| Demographic | Age, sex | Fundamental non-modifiable risk factors |
| Socioeconomic | Education, household income | Social determinants of health |
| Anthropometric | BMI, waist circumference | Indicators of adiposity and cardiometabolic risk |
| Lifestyle | Smoking, alcohol intake, activity, diet, sleep, stress | Modifiable behavioral risk factors |
| Medical history | Hypertension, diabetes | Baseline cardiometabolic risk |

#### 2.4.3. Model development and validation

Independent binary classification models were developed for each prediction horizon (5, 10 and 20 years). All analyses were performed independently for each prediction horizon using the same modelling workflow to ensure methodological consistency and facilitate direct comparison of predictive performance. Logistic regression was selected as the prediction algorithm because of its well-established performance in epidemiological research, probabilistic output and high degree of clinical interpretability, characteristics that facilitate subsequent integration into a clinical decision support system [24]. Rather than replacing clinical judgement, the objective of the model was to estimate individualized probabilities of long-term mortality as a decision-support tool for preventive nutritional assessment.

The analytical cohort was randomly divided into independent training (80%) and testing (20%) datasets. The training subset was used to estimate the model parameters, whereas the independent testing subset was reserved for evaluating predictive performance and generalizability. During model development, several iterative refinements were performed to improve model stability across prediction horizons, reduce the risk of overfitting, preserve clinical interpretability and ensure consistency between identified predictors and current epidemiological knowledge.

Model discrimination was evaluated using the area under the receiver operating characteristic curve (ROC-AUC), an established performance metric for binary classification problems with imbalanced outcomes [25].

#### 2.4.4. Feature importance analysis

Feature importance was assessed using permutation importance analysis to identify the variables contributing most to model performance [23]. The resulting rankings were subsequently used to support predictor interpretation and integration into the NutrIA CDSS.

### 2.5. Development of the NutrIA Clinical Decision Support System

#### 2.5.1. System architecture

The NutrIA Clinical Decision Support System (CDSS) was developed as a hybrid platform integrating supervised machine learning with knowledge-based clinical reasoning to support personalized nutritional assessment and preventive healthcare [6,13]. Rather than relying exclusively on statistical prediction, the system combines data-driven risk estimation with evidence-based clinical assessment, validated nutritional instruments and a rule-based recommendation engine to provide clinically interpretable outputs for healthcare professionals.

The architecture comprises two complementary components: (i) a machine learning model developed using NHANES data to estimate 5-, 10- and 20-year mortality risk, and (ii) a deterministic knowledge-based framework integrating validated clinical instruments, nutritional indicators and personalized recommendations. Together, these components provide transparent clinical decision support [26].

As illustrated in **Figure 2**, patient information is collected through an adaptive questionnaire and sequentially processed by interconnected modules responsible for clinical assessment, risk prediction, phenotype identification, dietary clustering and recommendation generation. The resulting outputs are integrated into an automated clinical report.

**Figure 2.**
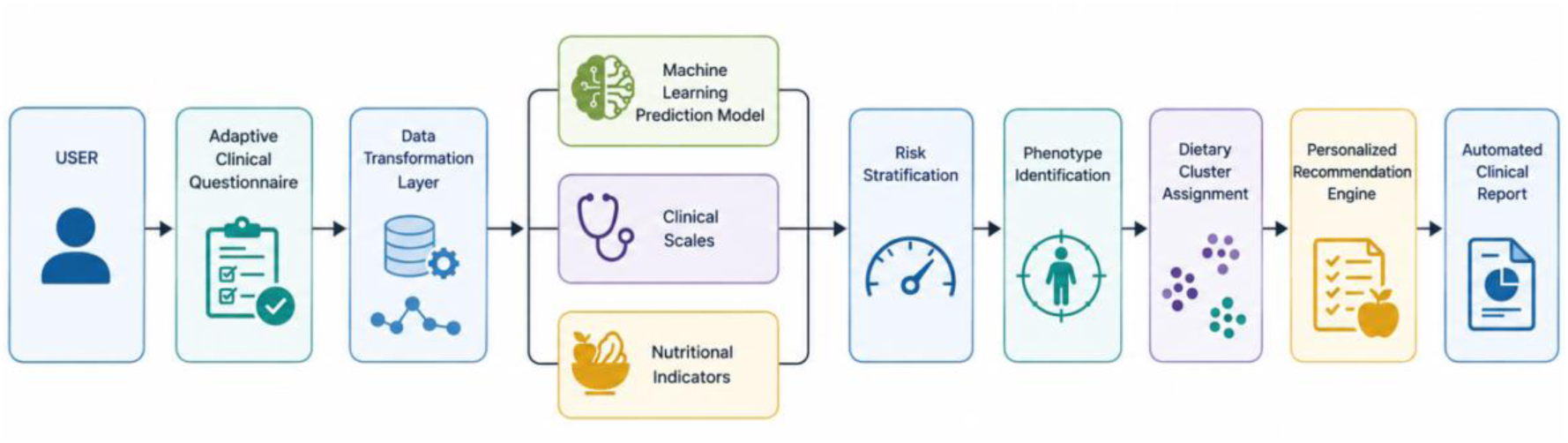
Overall architecture of the NutrIA Clinical Decision Support System.

#### 2.5.2. Web platform implementation

The NutrIA Clinical Decision Support System was implemented as a web-based platform to facilitate its use in routine nutritional assessment and preventive healthcare. The application provides an integrated digital environment in which users complete the adaptive clinical questionnaire, while healthcare professionals receive an automatically generated report containing individualized risk estimates, validated clinical assessments and personalized nutritional recommendations.

The platform follows a modular architecture in which patient responses are sequentially processed through interconnected components responsible for data transformation, clinical assessment, mortality risk prediction, risk stratification and personalized recommendation generation (**Figure 2**). This modular design facilitates future maintenance, scalability and integration of additional clinical algorithms.

The current implementation serves as a scalable research platform that allows incorporation of additional predictive models and clinical modules.

#### 2.5.3. Adaptive clinical questionnaire

The NutrIA Clinical Decision Support System incorporates an adaptive digital questionnaire specifically designed to collect the information required for both clinical assessment and machine learning-based risk prediction. Unlike conventional static questionnaires, the NutrIA questionnaire dynamically modifies its structure according to the user’s responses, allowing the assessment to focus on clinically relevant domains while minimizing respondent burden.

The questionnaire consists of a mandatory core section followed by multiple conditionally activated modules. The mandatory section collects essential demographic, anthropometric, socioeconomic and lifestyle information together with major cardiometabolic risk factors required for initial risk assessment. These variables also provide the minimum information necessary for calculating the mortality prediction models and several validated clinical instruments.

Following completion of the mandatory section, additional modules are activated automatically according to predefined clinical rules. Conditional activation is based on patient characteristics or previous responses, including age, body mass index, smoking status, alcohol consumption, physical activity, hypertension, diabetes, sleep quality, psychological stress and other clinically relevant conditions. This adaptive strategy enables comprehensive assessment of high-risk individuals while avoiding unnecessary questions for users without corresponding risk factors.

Beyond improving usability, the adaptive questionnaire automatically transforms patient-reported information into standardized variables compatible with both the machine learning model and the knowledge-based reasoning engine, thereby linking the predictive and deterministic components of the NutrIA CDSS.

The adaptive design reduces respondent burden while preserving the comprehensive information required for personalized nutritional assessment.

#### 2.5.4. Knowledge-based reasoning engine

The deterministic component of NutrIA is implemented through a knowledge-based reasoning engine that transforms patient-reported information into clinically interpretable assessments using structured decision rules derived from current clinical guidelines, validated screening instruments and nutritional expertise [13,26]. This component complements the machine learning prediction model by providing transparent clinical reasoning and facilitating the interpretation of individualized risk profiles.

The reasoning engine operates by sequentially processing the standardized variables generated from the adaptive questionnaire. Each variable is evaluated against predefined decision rules that determine the activation of validated clinical instruments, calculation of nutritional indicators, identification of clinical phenotypes and assignment of personalized recommendations. This rule-based approach ensures that all clinical outputs remain fully traceable, reproducible and directly linked to the corresponding patient responses.

The knowledge base consists of a structured repository of activation criteria, scoring algorithms, interpretation thresholds and recommendation rules. By integrating validated screening instruments, nutritional indicators and risk stratification algorithms, the reasoning engine generates a unified clinical interpretation that supports personalized preventive decision-making.

#### 2.5.5. Clinical assessment and validated instruments

The deterministic component of NutrIA incorporates a comprehensive set of validated clinical instruments together with internally developed nutritional indicators to support multidimensional assessment of nutritional status, lifestyle behaviours and cardiometabolic risk. Rather than relying exclusively on the mortality prediction model, the system performs a structured clinical evaluation across multiple health domains, providing interpretable information that complements probabilistic risk estimation.

Validated screening instruments were selected according to three principal criteria: widespread clinical use, scientific validation within their respective domains and applicability in preventive nutritional practice. The integrated instruments evaluate alcohol consumption, nicotine dependence, type 2 diabetes risk, cardiovascular risk, obstructive sleep apnoea, malnutrition, psychoactive substance use and fall risk, among other clinically relevant conditions [16,27–33]. Each instrument is calculated automatically using standardized scoring algorithms and internationally accepted interpretation thresholds.

In addition to validated instruments, NutrIA incorporates a series of internally developed clinical indicators designed to summarize complex behavioural and nutritional information into clinically interpretable measures. These indicators include multidimensional assessment of biological burden, functional reserve, preventive frailty, estimated biological age, post-event cardiovascular risk, familial cardiovascular risk and dietary quality. Although these indicators do not constitute diagnostic tools, they facilitate structured interpretation of modifiable risk factors and contribute to the personalization of preventive nutritional interventions.

All instruments and indicators are integrated within the knowledge-based reasoning engine, allowing simultaneous assessment of multiple clinical domains from a single patient questionnaire. The resulting outputs constitute the deterministic component of the NutrIA hybrid CDSS and provide the clinical context required to interpret the machine learning prediction model and support individualized decision-making.

#### 2.5.6. Risk stratification

Following completion of the clinical assessment, NutrIA performs an integrated risk stratification process that combines the outputs generated by the machine learning prediction model with the results of validated clinical instruments, internally developed nutritional indicators and structured clinical rules. This multidimensional approach enables the system to move beyond isolated risk estimation by providing a comprehensive interpretation of the patient’s preventive health profile.

The final risk profile integrates mortality prediction with multimorbidity, validated screening results, nutritional status and modifiable lifestyle factors, enabling the identification of priority areas for preventive intervention.

The resulting stratification is presented as an overall preventive risk category together with the principal determinants contributing to the assessment. This integrated profile constitutes the basis for the generation of personalized nutritional recommendations.

#### 2.5.7. Personalized recommendation engine

##### 2.5.7.1. Rule-based recommendation framework

The personalized recommendation engine constitutes the final decision-support component of the NutrIA hybrid Clinical Decision Support System. Its primary objective is to translate the outputs generated throughout the assessment process into individualized nutritional and lifestyle recommendations that are clinically interpretable and actionable. While the machine learning component estimates the probability of future mortality, the recommendation engine transforms this probabilistic information into practical preventive interventions adapted to each user’s clinical profile.

The recommendation framework is implemented as a deterministic rule-based system integrating demographic and anthropometric variables, patient-reported information, validated clinical instruments, nutritional indicators, mortality predictions and risk stratification results. These complementary data sources enable the automatic identification of modifiable risk factors and activation of personalized intervention pathways.

The knowledge base consists of a structured repository of activation criteria and recommendation rules linked to predefined clinical conditions, behavioural patterns and abnormal screening results. This transparent architecture ensures complete traceability between patient data, clinical interpretation and the generated recommendations.

Recommendations are prioritized according to their clinical relevance and organized into coherent intervention domains to facilitate interpretation by healthcare professionals and maintain consistency with evidence-based nutritional practice.

The recommendation engine supports, but does not replace, professional clinical judgement. Its deterministic structure facilitates future validation and continuous updating as new scientific evidence becomes available.

##### 2.5.7.2. Lifestyle recommendation module

The lifestyle recommendation module automatically generates individualized preventive recommendations aimed at improving modifiable health behaviours identified during the assessment process. This module represents the final deterministic layer of the NutrIA hybrid Clinical Decision Support System by translating the outputs of the clinical evaluation into structured and clinically interpretable guidance for both healthcare professionals and end users.

Recommendations are activated according to validated screening instruments, behavioural indicators, nutritional indices and clinically relevant findings identified during the assessment.

Lifestyle recommendations cover healthy eating, physical activity, sedentary behaviour, sleep hygiene, tobacco and alcohol use, medication adherence, self-care and fall prevention. Preventive screening recommendations are additionally generated according to age, sex and identified clinical risk factors.

The module also incorporates exploratory lifestyle indicators related to biological burden, functional reserve, behavioural risk and estimated biological age. These indicators provide complementary information for personalized counselling and should be considered exploratory pending further clinical validation.

Overall, the lifestyle recommendation module enables NutrIA to function not only as a predictive risk assessment system but also as a comprehensive preventive decision-support platform capable of transforming complex multidimensional clinical information into structured, personalized and actionable lifestyle recommendations.

##### 2.5.7.3. Personalized dietary recommendation module

The dietary recommendation module constitutes the nutrition-specific component of the NutrIA recommendation engine. It generates individualized dietary recommendations by integrating three complementary levels of personalization: estimation of individual nutritional requirements, dietary cluster assignment and clinical phenotyping. This multidimensional strategy simultaneously considers nutritional requirements, habitual dietary patterns and clinical priorities when generating dietary interventions.

###### Nutritional requirements

The first personalization layer estimates habitual dietary intake and individualized nutritional requirements. Habitual dietary intake is estimated from questionnaire responses using standardized food composition databases and serving-size conversion factors [34]. Individual energy and nutrient requirements are subsequently calculated using predictive equations for resting energy expenditure and self-reported physical activity [35–37]. Comparing habitual intake with individualized requirements enables identification of nutritional inadequacies and prioritization of dietary interventions.

###### Dietary clustering

The second personalization layer applies dietary clustering, an unsupervised machine learning approach [38], to assign each individual to the dietary pattern that best matches their eating behaviour. Clusters were predefined within age and weight-status strata to improve clinical applicability and identify priority dietary modifications toward healthier eating patterns.

###### Clinical phenotyping

The third personalization layer assigns a priority clinical phenotype using structured decision rules integrating anthropometric measurements, medical history, validated screening instruments and cardiometabolic risk factors. Phenotypes identify the principal nutritional condition requiring intervention, including obesity, metabolic syndrome, diabetes, elevated cardiovascular risk, gastrointestinal disorders and frailty.

Within the NutrIA framework, dietary clustering and clinical phenotyping perform complementary functions. The dietary cluster characterizes the individual’s habitual dietary pattern, whereas the clinical phenotype defines the principal therapeutic priority requiring nutritional intervention. Consequently, individuals presenting similar dietary patterns may receive different dietary recommendations according to their underlying clinical phenotype, while patients sharing the same phenotype may receive different nutritional priorities depending on their baseline dietary habits.

Final dietary recommendations are generated through the integration of these three personalization layers. This approach enables NutrIA to produce individualized nutritional guidance that considers dietary adequacy, habitual eating behaviour and clinical priorities within a unified decision-support framework.

#### 2.5.8. Automated clinical report generation

The final output of NutrIA is an automatically generated clinical report that integrates all assessment results into a structured and clinically interpretable document.

The report is generated automatically following completion of the adaptive questionnaire. Outputs from the machine learning model, validated clinical instruments, nutritional indicators, risk stratification, dietary clustering, clinical phenotyping and the recommendation engine are consolidated into a unified report without additional user input. This automated workflow ensures consistent and standardized presentation of patient information. An example of the automatically generated clinical report is presented in **Figure 3**.

**Figure 3.**
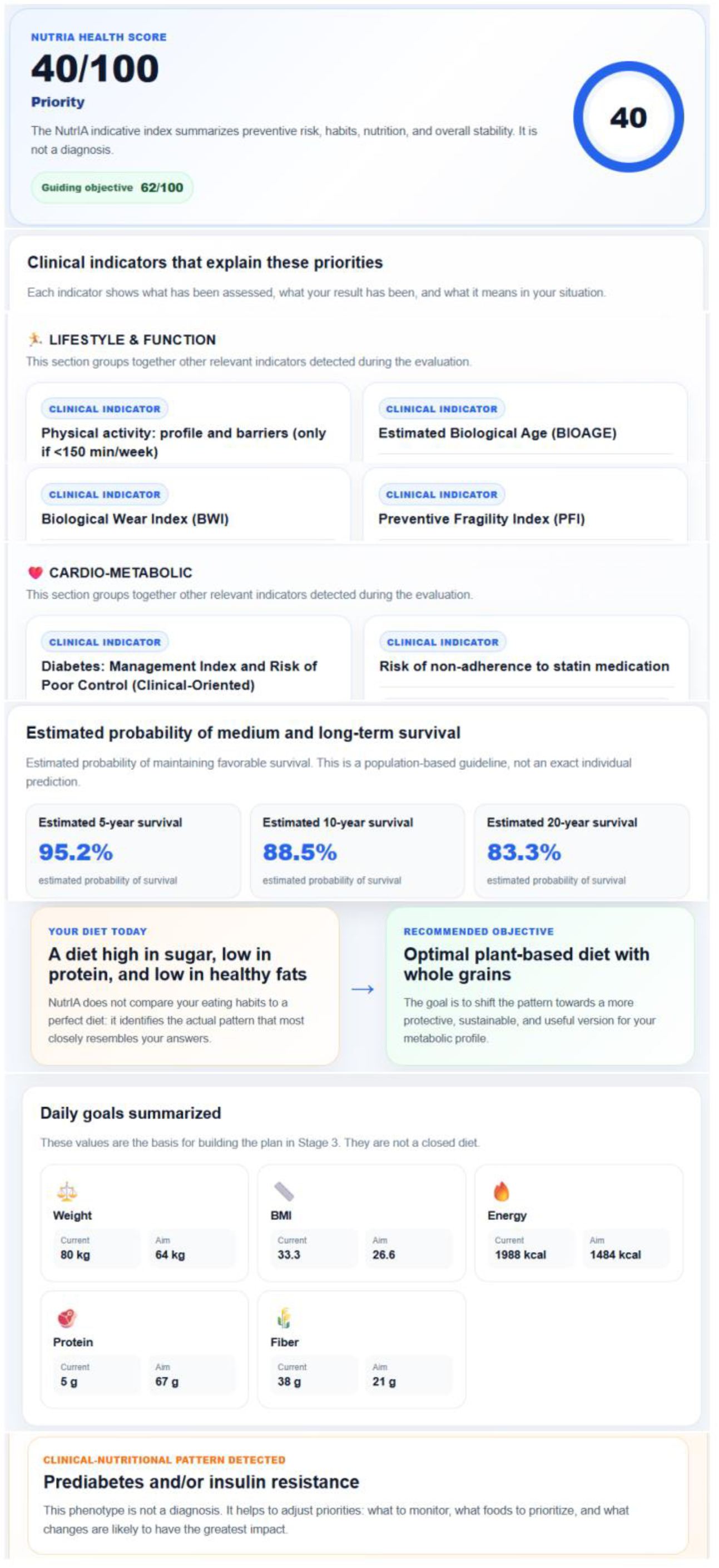
Reduced example of the automated clinical report generated by NutrIA.

The report is organized into clinically meaningful sections summarizing demographic and anthropometric characteristics, estimated mortality risk, validated clinical assessments, nutritional indicators, risk profile, dietary cluster, clinical phenotype, estimated nutritional requirements and personalized recommendations. This structure facilitates rapid identification of priority preventive interventions while preserving traceability between patient responses and system outputs.

The automated report is intended to function as a clinical decision-support resource rather than a diagnostic document. Consequently, all information generated by NutrIA should be interpreted by qualified healthcare professionals within the context of a comprehensive nutritional assessment. By integrating predictive analytics with knowledge-based reasoning, the report translates complex multidimensional data into clinically actionable information for preventive nutritional practice.

## 3. Results

### 3.1. Final NutrIA architecture

The final implementation of NutrIA resulted in a hybrid Clinical Decision Support System integrating machine learning prediction, knowledge-based clinical reasoning and automated personalized nutrition support within a unified digital platform. The implemented architecture combines an adaptive clinical questionnaire, a mortality prediction model, validated screening instruments, internally developed nutritional indicators, multidimensional risk stratification and an automated recommendation engine into a single workflow capable of supporting preventive nutritional assessment.

The completed system consists of several interoperable modules that operate sequentially during the assessment process. Initially, users complete an adaptive questionnaire in which conditional logic dynamically activates additional questions according to previous responses. The collected information is subsequently processed to generate standardized clinical variables, validated screening scores, derived nutritional indicators and the predictors required by the machine learning model. These outputs are then integrated into a rule-based reasoning engine that performs multidimensional risk stratification, dietary cluster assignment and clinical phenotype identification before generating individualized lifestyle and dietary recommendations. Finally, all results are consolidated into an automatically generated clinical report designed to facilitate interpretation by healthcare professionals.

The final implementation incorporates **39 clinical assessment scales**, multiple internally developed nutritional indicators, automated estimation of individualized nutritional requirements, dietary pattern classification through clustering techniques, structured clinical phenotyping and a transparent rule-based recommendation engine. These components complement the mortality prediction model by transforming probabilistic risk estimates into clinically interpretable preventive recommendations, extending the functionality of the system beyond risk prediction alone.

### 3.2. Predictive performance

The machine learning component of NutrIA demonstrated consistently high discriminative performance for predicting all-cause mortality across the three prediction horizons evaluated. The 5-year model achieved a receiver operating characteristic area under the curve (ROC-AUC) of **0.894**, while the 10-year and 20-year models achieved ROC-AUC values of **0.914** and **0.923**, respectively, indicating excellent discrimination across all prediction horizons.

The prevalence of mortality events increased with longer follow-up periods, from **6.18%** at 5 years to **11.38%** at 10 years and **15.64%** at 20 years, reflecting the expected accumulation of events over time. Despite these differences in event prevalence, predictive performance remained consistently high across all evaluated models.

**Table 4** summarizes the predictive performance obtained for each prediction horizon.

**Table 4.** Predictive performance of the mortality prediction models.

| Prediction horizon | Event prevalence (%) | ROC-AUC |
| --- | --- | --- |
| 5 years | 6.18 | 0.894 |
| 10 years | 11.38 | 0.914 |
| 20 years | 15.64 | 0.923 |

### 3.3. Feature importance analysis

Feature importance analysis identified age as the strongest predictor of mortality across all prediction horizons. Although its relative contribution gradually decreased with increasing follow-up duration, it remained the dominant predictor in every model.

Anthropometric variables, particularly indicators of body composition and central adiposity, demonstrated substantial predictive importance, especially for short-term mortality prediction. In contrast, self-reported chronic diseases, cardiometabolic conditions and socioeconomic variables became progressively more influential in the 10- and 20-year prediction models. Lifestyle-related variables, including smoking, alcohol consumption, dietary quality and physical activity, consistently contributed to model discrimination across all prediction horizons.

The observed agreement between permutation feature importance and established epidemiological evidence supports the biological plausibility and clinical interpretability of the developed prediction models.

### 3.4. Clinical workflow

The implemented NutrIA workflow follows a structured sequence that transforms patient-provided information into personalized clinical decision support through the integration of machine learning prediction and deterministic clinical reasoning.

The workflow begins with completion of the adaptive questionnaire, where mandatory information is collected for all users and additional question blocks are dynamically activated according to previous responses. This adaptive strategy allows comprehensive clinical characterization while minimizing unnecessary data collection and reducing respondent burden. Upon completion of the questionnaire, all responses are automatically standardized and converted into structured variables suitable for computational analysis.

The processed information is subsequently analysed by multiple computational modules operating within the same decision-support framework. The machine learning component estimates individual probabilities of 5-, 10- and 20-year all-cause mortality, while the deterministic reasoning engine simultaneously calculates validated clinical assessment scales, internally developed nutritional indicators, integrated risk stratification, dietary cluster assignment and priority clinical phenotype. The outputs are integrated to generate an individualized preventive assessment.

Finally, the outputs produced by all modules are integrated by the recommendation engine, which generates personalized lifestyle and dietary interventions tailored to the individual’s clinical profile, nutritional requirements and behavioural characteristics. The complete assessment is presented through an automatically generated clinical report that summarizes risk estimates, principal findings and recommended preventive actions in a standardized format suitable for clinical interpretation.

### 3.5. Clinical use case

To illustrate the practical application of NutrIA, a fully hypothetical clinical profile was processed through the complete Clinical Decision Support System. This simulated example does not correspond to any real patient or study participant. The profile represented a woman in the 50–60-year age range with anthropometric characteristics compatible with overweight. She completed the adaptive questionnaire and underwent the full automated assessment workflow. Following questionnaire completion, all computational modules were executed automatically without requiring additional user input, resulting in an integrated clinical report summarizing the simulated nutritional and preventive health profile (**Figure 4**).

**Figure 4.**
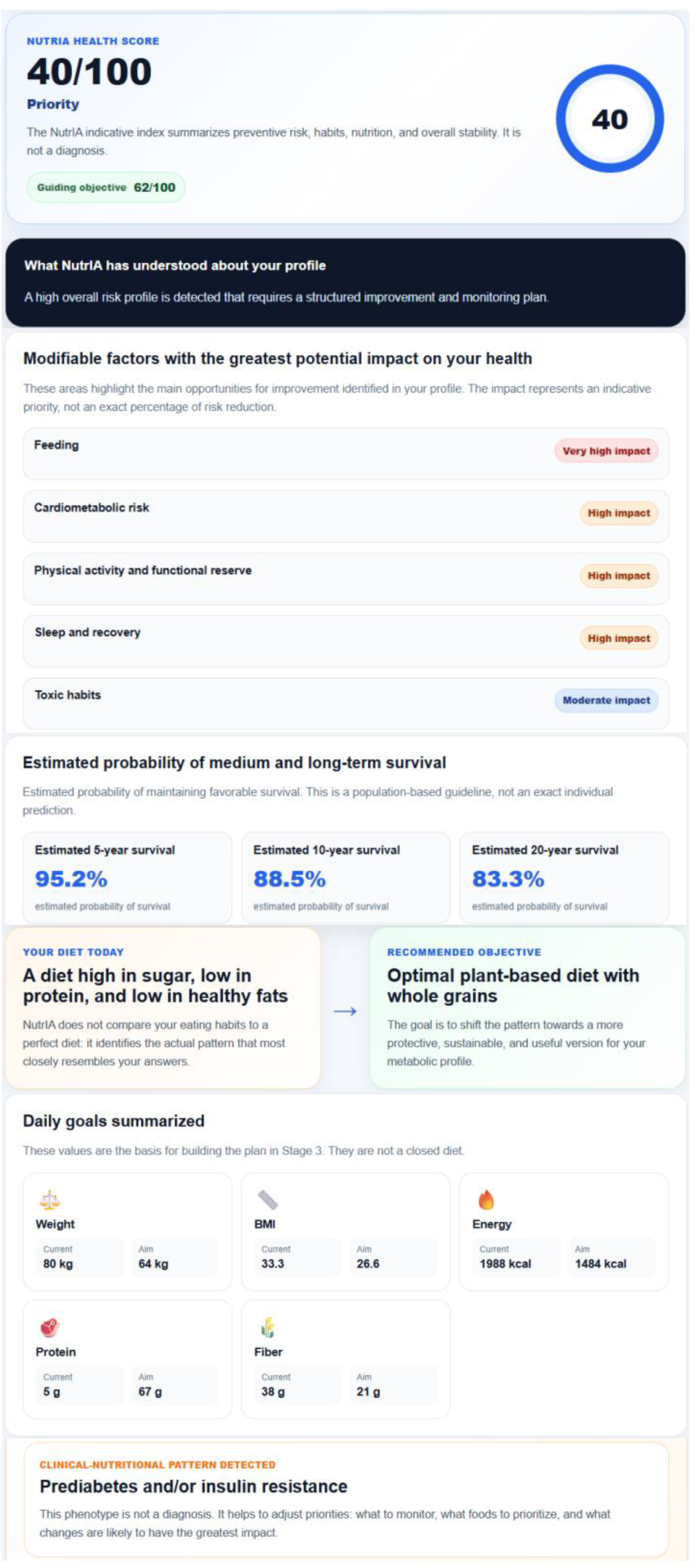
Reduced clinical report of a hypothetical simulated use case.

The integrated assessment identified an overall high preventive risk profile characterized by severe sedentary behaviour, elevated cardiometabolic risk, reduced functional reserve, accelerated biological ageing and suboptimal dietary quality. The adaptive questionnaire activated multiple validated clinical instruments, including FINDRISC, STOP-Bang and the Healthy Eating Index, together with several internally developed nutritional indicators evaluating biological ageing, preventive frailty, functional reserve and metabolic risk. Collectively, these findings enabled comprehensive characterization of the patient’s nutritional, metabolic and behavioural status.

The knowledge-based reasoning engine subsequently integrated these results with the mortality prediction model to generate a unified clinical interpretation. In addition to estimating individualized nutritional requirements, the system assigned the patient to a predefined dietary cluster representing the habitual eating pattern and identified the principal clinical phenotypes requiring nutritional intervention, including metabolic syndrome, cardiovascular risk, dyslipidaemia, abdominal obesity and insulin resistance. These complementary analyses established the priorities used by the recommendation engine to personalize dietary and lifestyle interventions.

Based on the integrated assessment, NutrIA automatically generated individualized recommendations targeting physical activity, dietary quality, weight management, cardiometabolic prevention and participation in age- and sex-appropriate screening programmes. Nutritional guidance included estimated energy and nutrient requirements, identification of dietary inadequacies, recommended modifications to the habitual dietary pattern and phenotype-specific nutritional strategies designed to improve long-term metabolic health.

Finally, the recommendation engine simulated the potential impact of adopting the proposed dietary pattern within the predictive framework implemented in NutrIA. For this illustrative case, the model estimated a substantial reduction in long-term mortality risk following implementation of the personalized nutritional strategy. These values represent model-derived predictions generated by the integrated decision-support framework and are intended to illustrate the potential clinical application of the system rather than observed patient outcomes.

## 4. Discussion

### 4.1. Principal findings

The present study describes the development and internal validation of NutrIA, a hybrid Clinical Decision Support System that integrates machine learning, validated clinical assessment instruments and transparent rule-based reasoning to support personalized nutrition in preventive healthcare [6,13]. Unlike conventional nutritional software, which primarily focuses on dietary planning, or isolated prediction models that estimate a single clinical outcome, NutrIA combines predictive analytics with multidimensional nutritional assessment and automated recommendation generation within a unified digital platform [7,26,39].

The machine learning component demonstrated excellent predictive performance for predicting 5-, 10- and 20-year all-cause mortality, with ROC-AUC values ranging from 0.894 to 0.923. However, the principal contribution of this work extends beyond predictive performance.

An additional contribution of this work is the implementation of an adaptive clinical questionnaire capable of reducing unnecessary data collection while maintaining the information required for multidimensional nutritional assessment. The integration of adaptive assessment, predictive modelling and automated report generation allows NutrIA to support the complete preventive nutrition workflow rather than functioning as a standalone prediction algorithm.

Overall, the proposed architecture demonstrates the feasibility of combining data-driven prediction models with explicit clinical knowledge within a single CDSS, providing healthcare professionals with structured, interpretable and personalized information to facilitate preventive nutritional decision-making [13,26].

### 4.2. Comparison with previous studies

Recent advances in artificial intelligence have substantially improved the prediction of chronic diseases and long-term health outcomes through the application of machine learning to large epidemiological datasets [10,12]. Several predictive models have reported excellent discrimination for cardiovascular disease, type 2 diabetes and all-cause mortality using approaches ranging from traditional statistical models to more complex machine learning algorithms [40–43]. Well-established clinical tools such as QRISK3 and the Framingham Risk Score have demonstrated considerable value for cardiovascular risk prediction [15,44], while more recent machine learning studies have further improved predictive accuracy by incorporating multidimensional health data [7,12,40]. However, these approaches generally function as standalone prediction models, providing probability estimates that require subsequent interpretation by healthcare professionals rather than supporting the complete clinical decision-making process.

Clinical Decision Support Systems (CDSSs) have progressively evolved from simple rule-based applications towards hybrid systems combining predictive analytics with evidence-based clinical reasoning [6,13]. Previous studies have highlighted the potential of explainable artificial intelligence and transparent decision-support architectures to improve clinician confidence, facilitate implementation and support evidence-based healthcare [6,13, 26]. Nevertheless, most existing CDSSs remain focused on specific diseases or individual clinical tasks and rarely integrate predictive modelling, validated nutritional assessment, automated interpretation and personalized preventive recommendations within a single interoperable platform.

Within clinical nutrition, commercially available software platforms such as Nutrium, Dietopro and EasyDiet have substantially improved dietary management, nutritional follow-up and patient monitoring. However, these systems primarily function as nutritional management tools and generally do not incorporate predictive machine learning models, adaptive clinical questionnaires, dietary clustering, clinical phenotyping or multidimensional risk stratification to support preventive clinical decision-making. Consequently, nutritional recommendations continue to depend largely on manual interpretation by healthcare professionals rather than on the integration of predictive analytics with structured clinical reasoning.

To our knowledge, NutrIA is among the first web-based CDSS specifically developed for preventive nutrition that combines machine learning prediction, adaptive clinical assessment, validated screening instruments, dietary clustering, clinical phenotyping and transparent rule-based reasoning within a single interoperable platform. Rather than functioning as an isolated prediction model or a conventional dietary management application, NutrIA transforms probabilistic risk estimates into clinically interpretable recommendations through the simultaneous integration of multiple complementary sources of nutritional and clinical information [14,19]. This hybrid architecture allows healthcare professionals to understand not only the estimated level of risk but also the clinical rationale underlying each recommendation generated by the system.

Finally, the present study extends previous research by demonstrating the technical feasibility of integrating machine learning prediction, structured clinical reasoning and automated report generation into a fully implemented web-based CDSS [7]. Although external validation and prospective clinical evaluation remain necessary before routine implementation, the proposed architecture represents an important step towards clinically interpretable artificial intelligence capable of supporting personalized preventive nutrition in real-world healthcare settings [14,19].

### 4.3. Clinical implications

The increasing prevalence of chronic non-communicable diseases highlights the need for clinical tools capable of supporting early risk identification and personalized preventive interventions. In this context, NutrIA was designed not to replace clinical judgment but to provide healthcare professionals with an integrated decision-support system that facilitates evidence-based nutritional assessment and individualized dietary planning [6,13].

By combining machine learning predictions with validated screening instruments, dietary pattern analysis, clinical phenotyping and transparent rule-based reasoning, the proposed system enables clinicians to obtain a comprehensive overview of each patient’s nutritional and preventive health status through a single automated workflow [26]. This integration may reduce the time required for data interpretation, improve consistency in nutritional assessments and facilitate the identification of modifiable risk factors that could otherwise remain unnoticed during routine consultations.

The adaptive questionnaire incorporated into NutrIA also has practical implications for clinical usability. By dynamically selecting only the questions that are relevant for each individual, the system minimizes respondent burden while maintaining a multidimensional evaluation of nutritional, behavioural and clinical domains. This approach may improve patient engagement and increase the feasibility of implementing comprehensive nutritional assessments in busy healthcare settings.

From a professional perspective, the automated generation of structured clinical reports may contribute to standardizing nutritional evaluations across different healthcare environments while preserving transparency in the decision-making process. Unlike black-box predictive models, NutrIA presents the rationale underlying its recommendations, allowing healthcare professionals to critically interpret the generated outputs and incorporate them into their own clinical reasoning [26]. This characteristic is particularly important in preventive nutrition, where individualized interventions frequently require the integration of multiple sources of clinical information.

Although the present study focused on internal development and validation, the proposed architecture could potentially be adapted to multiple healthcare contexts, including primary care, hospital nutrition units, preventive medicine services and telehealth programmes. Consequently, hybrid Clinical Decision Support Systems such as NutrIA may represent a promising strategy for improving the efficiency, standardization and personalization of preventive nutritional care.

### 4.4. Strengths and limitations

The present study has several strengths. First, NutrIA was developed using a large nationally representative dataset comprising 59,064 participants from NHANES linked to mortality records, providing a robust foundation for predictive modelling and reducing the risk of overfitting associated with smaller datasets. Second, rather than focusing exclusively on predictive performance, the proposed system integrates machine learning with validated screening instruments, dietary pattern analysis, clinical phenotyping and transparent rule-based reasoning within a single Clinical Decision Support System. This multidimensional architecture enhances the interpretability and practical applicability of the generated recommendations. Finally, the implementation of a fully functional web-based platform demonstrates the technical feasibility of translating predictive models into a usable clinical decision-support tool.

Nevertheless, several limitations should be acknowledged. First, the predictive model was developed and internally validated using NHANES data, and external validation in independent populations is required before widespread clinical implementation [45,46]. Population characteristics, healthcare systems and dietary habits may differ across countries, potentially affecting the generalizability of the model. Second, although the recommendation engine is based on evidence-informed clinical rules and validated assessment instruments, its clinical effectiveness has not yet been evaluated through prospective intervention studies or randomized clinical trials [13]. Third, the current version of NutrIA was designed to support preventive nutritional assessment in adults and may require adaptation before being applied to specific populations such as children, pregnant women or patients with highly specialized nutritional needs.

In addition, the current study evaluated the technical performance of the platform but did not assess usability from the perspective of healthcare professionals or patients.

Despite these limitations, the proposed hybrid architecture provides a flexible framework that can be progressively refined as new datasets, predictive models and clinical evidence become available. Future external validation studies and prospective evaluations will be essential to determine its clinical impact, usability and effectiveness in routine healthcare practice.

### 4.5. Future research

Future research should focus on the external validation of NutrIA in independent populations from different geographical regions and healthcare settings to evaluate its generalizability and robustness across diverse demographic and clinical contexts. Prospective clinical studies are also required to determine whether the use of the system improves nutritional assessment, clinical decision-making, patient adherence and health outcomes compared with conventional nutritional care [45,46].

Further development of the platform may include the incorporation of additional data sources, such as electronic health records, wearable devices and continuous lifestyle monitoring systems, allowing dynamic updates of individual risk profiles and nutritional recommendations [7,39]. Future versions could also integrate more advanced machine learning approaches, including multimodal and longitudinal models, while maintaining the transparency and interpretability required for routine clinical practice [14]. In addition, expanding the system to specific populations, such as pediatric, geriatric or oncology patients, may increase its applicability across different areas of clinical nutrition [47].

Future work should also evaluate the incorporation of large language models to facilitate conversational nutritional counselling while maintaining transparency and clinical supervision [48].

Finally, future work should evaluate the usability, acceptance and cost-effectiveness of NutrIA in real-world healthcare environments. Such studies will be essential to determine the clinical value of hybrid Clinical Decision Support Systems and to facilitate their integration into routine preventive healthcare services [13,14].

## 5. Conclusions

This study presents NutrIA, a hybrid web-based Clinical Decision Support System that integrates machine learning, validated clinical assessment instruments, dietary clustering, clinical phenotyping and transparent rule-based reasoning to support personalized preventive nutrition. Unlike conventional nutritional software or standalone prediction models, NutrIA combines predictive analytics with multidimensional clinical assessment and automated recommendation generation within a single interoperable platform.

The proposed system demonstrated excellent predictive performance for 5-, 10- and 20-year all-cause mortality while illustrating the feasibility of translating machine learning outputs into clinically interpretable decision support through the integration of adaptive assessment, evidence-based clinical reasoning and automated report generation. This hybrid architecture enables healthcare professionals to obtain individualized nutritional evaluations and personalized preventive recommendations using a standardized and transparent workflow.

Although external validation and prospective clinical evaluation are required before routine implementation, NutrIA provides a proof of concept for the integration of predictive modelling, explainable artificial intelligence and structured clinical reasoning within a single CDSS for preventive nutrition. To our knowledge, this represents one of the first web-based platforms specifically designed to combine these complementary components into an integrated framework for personalized nutritional care. The proposed approach may contribute to the future development of clinically interpretable artificial intelligence capable of supporting evidence-based preventive nutrition in routine healthcare practice.

## Acknowledgements

The authors would like to express their sincere gratitude to the Universidad Internacional de La Rioja (UNIR) for providing the academic environment in which this research was developed.

The authors also acknowledge the Centro de Investigación en Atención Primaria (CIAP), Miguel Hernández University of Elche, and the Parque Científico de la Universidad Miguel Hernández (PCUMH) for promoting innovation and knowledge transfer in digital health and preventive medicine.

The authors gratefully acknowledge Dana Assist for its technical collaboration, software development expertise and continuous support during the design, implementation and deployment of the NutrIA web-based Clinical Decision Support System.

## Data Availability Statement

The datasets analyzed during the current study are publicly available through the National Health and Nutrition Examination Survey (NHANES) program of the National Center for Health Statistics (NCHS) [20]. Additional data generated and analyzed during this study are available from the corresponding author upon reasonable request.

## Code Availability Statement

The source code, knowledge base and proprietary algorithms underlying the NutrIA platform are not publicly available because they form part of an actively developed software project.

A demonstration version of the NutrIA web platform is publicly available at: https://nutria-web-60434848629.europe-west1.run.app/app

Additional methodological information may be obtained from the corresponding author upon reasonable request. NutrIA is currently under active development by Dana Assist.

## Declaration of Generative AI

During the preparation of this manuscript, the authors used generative artificial intelligence (ChatGPT, OpenAI) to assist with language editing, text refinement and manuscript organization. All scientific concepts, study design, methodology, software development, data analysis, interpretation of results and final editorial decisions were performed, critically reviewed and approved by the authors, who accept full responsibility for the content of this manuscript.

## CRediT Author Contributions

Bohdan Isaiev: Conceptualization, Methodology, Software, Formal analysis, Validation, Supervision, Project administration, Funding acquisition (if applicable), Writing – review & editing.

Iryna Stukalova: Conceptualization, Investigation, Methodology, Data curation, Validation, Visualization, Writing – original draft, Writing – review & editing.

## Ethics Statement

This study was based exclusively on publicly available, de-identified data from the National Health and Nutrition Examination Survey (NHANES) [20]. Therefore, no additional ethical approval or informed consent was required for the present secondary data analysis.

